# Clonal Hematopoiesis and Risk of Stroke: Evidence from Over 800,000 Individuals Across Three Cohorts

**DOI:** 10.1101/2025.10.22.25338594

**Authors:** Kun Zhao, Yash Pershad, Liying Xue, Caitlyn Vlasschaert, Robert W Corty, J Brett Heimlich, Charles Kooperberg, Alexander P Reiner, Alexander G Bick

**Author notes:** Correspondence to: Dr. Bick, Address: 550 Robinson Research Building, Nashville, TN 37232. These authors contributed equally to this work (Kun Zhao, Yash Pershad).

## Abstract

Clonal hematopoiesis of indeterminate potential (CHIP) is a common age-related condition that increases risk for cardiovascular disease. However, its relationship with stroke remains uncertain: some studies have reported a significant association between CHIP and stroke risk, while others, including large biobank analyses, found no association after adjustment. To resolve these conflicting findings, we analyzed genomic and clinical data from 800,160 participants with genetic sequencing and medical records across the Vanderbilt BioVU, NIH *AllofUs*, and UK Biobank. Stroke events were identified and classified as ischemic or hemorrhagic using ICD codes. Results from the three cohorts were meta-analyzed with previously published results. Subgroup analyses were conducted by driver gene, clone size, sex and menopausal status. In addition, genetically predicted levels of 27 plasma cytokines were assessed as potential modifiers of CHIP-associated stroke risk. CHIP was associated with increased risk of incident stroke in each cohort and in the meta-analysis (HR = 1.20, 95% CI 1.13–1.27; *P* = 2.21 × 10^⁻10^). This association was observed for both ischemic (HR = 1.18) and hemorrhagic (HR = 1.30) stroke subtypes. Gene-specific analyses showed strong associations for *JAK2* (HR = 2.46) and *TET2* (HR = 1.40). *DNMT3A* demonstrated weak but significant associations (HR = 1.11). CHIP was associated with stroke risk in both sexes; however, among women, the association was evident in postmenopausal (HR = 1.49, 95% CI 1.16–1.92; *P* = 1.91 × 10^⁻3^) but not in premenopausal participants (HR = 0.70, 95% CI 0.36–1.43, *P* = 0.33). Among participants with CHIP, but not among participants without CHIP, genetically predicted levels of IL-1RAP were predictive of risk for stroke, suggesting IL-1RAP as a modifier of the CHIP-associated risk for stroke. Collectively, this large-scale, multi-cohort study establishes CHIP as an important determinant of incident stroke risk and IL-1-mediated inflammation as a targetable pathway to reduce this risk.

## Introduction

Stroke is one of the most devastating non-communicable diseases, ranking as the second leading cause of death worldwide (∼7 million annually) and the third leading cause of death and disability combined, accounting for more than 160 million disability-adjusted life years.^1–3^ Stroke occurs more frequently in men and the elderly, while women face disproportionately higher risks of stroke-related mortality and long-term disability.^4–6^ Ischemic and hemorrhagic strokes represent distinct entities, with divergent pathophysiological mechanisms and risk factors.^3^ While ischemic stroke is primarily driven by atherosclerosis and thrombosis, hemorrhagic stroke is more strongly linked to hypertension, vascular fragility, and coagulation abnormalities.^7^ Inflammation plays a central role in the pathogenesis and outcomes of both types of stroke.^8,9^ Cytokines represent key mediators in this process.^10–12^ Pro-inflammatory cytokines, such as IL-1^13–15^ and IL-6^16–18^, have been shown to exacerbate ischemic brain injury by activating immune cells, disrupting the blood–brain barrier, and amplifying secondary tissue damage. Conversely, anti-inflammatory cytokines like IL-10 may mitigate adverse inflammatory responses and improve stroke prognosis.^19,20^ While stroke is influenced by multiple risk factors, including lifestyle, vascular comorbidities and menopause, genetic susceptibility has also been firmly established as an important determinant of overall stroke risk.^21–23^ Beyond inherited germline variation, emerging evidence suggests that acquired somatic mutations may also contribute to stroke susceptibility.^24,25^

Clonal hematopoiesis of indeterminate potential (CHIP) is the presence of an expanded somatic blood cell clone in individuals without other hematologic abnormalities, most commonly involving mutations in *DNMT3A* and *TET2*.^26,27^ CHIP is observed in >10% of individuals aged 70 years or older, is strongly associated with atherosclerotic cardiovascular disease.^28–31^ The association between CHIP and stroke was first reported by Jaiswal et al.,^24^ who demonstrated a 2.6-fold increased risk of ischemic stroke among CHIP carriers in a cohort of 3,913 individuals. Subsequently, in a meta-analysis of more than 80,000 participants across 8 prospective studies, Bhattacharya et al.^25^ reported a statistically significant increase in total stroke risk (hazard ratio [HR] =1.14, [95% CI, 1.03-1.27]; *P*=0.01); the risk associated with CHIP was strongest for hemorrhagic and small-vessel ischemic stroke subtypes. More recent large-scale analyses of the UK Biobank have yielded conflicting results. For example, Kessler et al.^32^ reported that only *JAK2*-CHIP was associated with the prevalence of overall stroke and ischemic stroke, while Kar et al.^33^ found that CHIP was not significantly associated with overall stroke incidence (HR range 0.90–1.72; *P* > 0.09). A recent study further suggest that CHIP is associated with increased white matter lesion burden, and a proinflammatory profile in ischemic stroke patients, and that mutations such as *TET2* confer higher risks of recurrent vascular events and mortality after ischemic stroke.^34^ Recent multi-omics and Mendelian randomization studies have suggested causal links between CHIP and ischemic stroke via inflammation.^35^

These divergent findings underscore the need to clarify the relationship between CHIP and stroke at-scale. To address this gap, we analyzed CHIP and stroke outcomes in more than 800,000 individuals from three large independent cohorts and further incorporated published results from the Women’s Health Initiative into meta-analyses. We aimed to (1) systematically evaluate the association between CHIP and incident stroke, in aggregate and by major subtype (i.e., ischemic and hemorrhagic), (2) explore whether the relationship varies by mutated gene, clone size, sex or menopausal status, and (3) investigate potential targetable mechanisms via genetically predicted cytokines.

## Methods

### Study population and design

This study utilized data from three large-scale cohorts: the Vanderbilt’s BioVU biobank, NIH *AllofUs* (AoU) Research Program, and the UK Biobank (UKB). Participants with available whole genome or exome sequencing data were initially considered (BioVU: 245,540; AoU: 275,679; UKB: 488,175). We excluded individuals with a documented history of stroke prior to enrollment, a history of hematologic malignancy or other non-neoplastic clonal disorders, lack of follow-up information, or missing key covariates. As CHIP is highly uncommon in younger individuals, to minimize potential technical confounding, we excluded participants with baseline age <40, thereby including 800,160 participants across three cohorts (**Figure 1**). Although UKB results have been reported in prior publications,^25,32,33^ our analyses leveraged updated genomic data, which included several hundred thousand additional participants not previously examined as well as updated outcomes data including 18,553 additional events since the last analysis (N= 4,321).^33^

**Figure 1.**
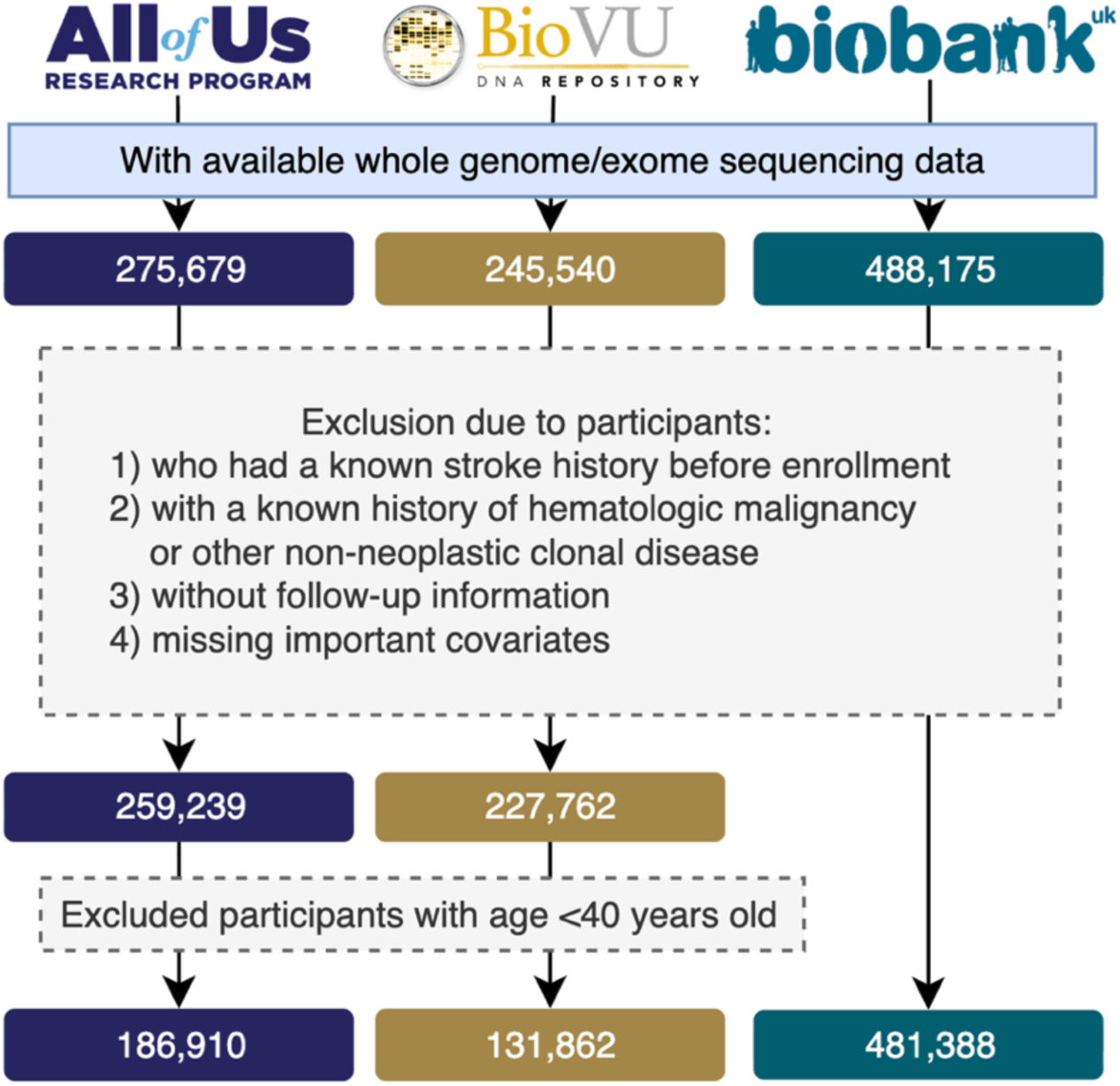
Study population flowchart across three cohorts. Flow diagram showing participant selection in the *All of Us* Research Program, Vanderbilt BioVU, and UK Biobank cohorts. Individuals were excluded if they had (1) a known history of stroke before enrollment, (2) a history of hematologic malignancy or other non-neoplastic clonal disease, (3) missing follow-up information, or (4) missing key covariates. Participants younger than 40 years were further excluded. The final analytic cohorts included 186,910 participants from *All of Us*, 131,862 from BioVU, and 481,388 from UK Biobank

### Clonal hematopoiesis of indeterminate potential variant calls

Putative somatic SNVs and short indels were called with GATK *Mutect2* and filtered according to previously established criteria among the three cohorts as we have previously described.^36,37^ Briefly, 74 canonical CHIP genes were screened for potential CHIP mutations using the *Mutect2* somatic variant caller **(Supplementary Table 1)**.^28^ Variants included in the preliminary dataset met the following criteria: presence in a pre-established list of candidate CHIP variants, total sequencing depth ≥ 20, alternate allele read depth count ≥ 5, and representation in both sequencing directions (i.e., F1R2 ≥ 1 and F2R1 ≥ 1). CHIP mutations were defined as those with a variant allele fraction (VAF) ≥ 0.02. CHIP detection across all cohorts was derived from whole genome/exome sequencing data, using a consistent detection method and the same canonical CHIP driver genes list, which specifies candidate missense and indel variants for each gene, as well as a set of genes in which truncating and splice site variants may be considered.

### Study outcome and covariates

The primary outcome was incident stroke, with additional analyses performed for the subtypes of ischemic stroke and hemorrhagic stroke. Outcomes were ascertained using the International Classification of Diseases (ICD) codes (ICD-9 and ICD-10) and harmonized across the three cohorts to ensure consistent definitions **(Supplementary Table 2)**. Covariates included baseline age, sex, smoking status, principal components of ancestry (PC1–PC5), low-density lipoprotein cholesterol (LDL-C), hypertension, body mass index (BMI), and type 2 diabetes. Current cigarette smoking was categorized as current smoker versus non-smoker. BMI (kg/m²) was calculated from measured height and weight at the baseline visit. Hypertension was defined as a diagnosis based on ICD codes prior to baseline or elevated blood pressure at baseline (systolic or diastolic in mmHg). Type 2 diabetes was defined as a diagnosis based on ICD codes prior to baseline or an elevated baseline fasting glucose (mg/dL). LDL-C (mmol/L) levels were obtained from baseline laboratory assessments. Given the well-established association between atrial fibrillation (AF) and stroke, we additionally adjusted for baseline AF status in sensitivity analyses to ensure the robustness of the observed associations.

### Statistical analysis

Descriptive statistics were used to summarize baseline characteristics. Continuous variables were reported as mean (SD), and categorical variables as counts and percentages. The association between CHIP and incident stroke was assessed using age-scaled Cox proportional hazards models, adjusting for the covariates described above. Left truncation was accounted for by defining each participant’s entry age as age at baseline and exit age as age at event or censoring. The outcome was defined as the first recorded stroke event. Participants were followed from baseline until the earliest occurrence of stroke or end of follow-up. For subtype-specific analyses, only the first ischemic or hemorrhagic stroke was considered as an event of interest. To obtain pooled estimates, we combined results from these three cohorts with those from the Bhattacharya study,^25^ restricting to cohorts in which the number of CHIP carriers with stroke exceeded 100 for meta-analysis (i.e., the Women’s Health Initiative, or WHI, with CHIP calls ascertained from whole-genome sequencing as in our studies). Inverse variance–weighted meta-analysis was performed under either a fixed- or random-effects framework, selected according to the degree of heterogeneity (I² statistic). Additional analyses were conducted stratified by VAF of CHIP mutations (≥2% and ≥10% versus no mutation), sex, and menopausal status for women (menopausal data was only available in UKB). To investigate whether menopausal status altered the association between CHIP and stroke in women, we conducted a sensitivity analysis using age-matched groups. Specifically, we applied 1:1 nearest-neighbor matching on baseline age between postmenopausal and premenopausal women using the *MatchIt* package in R. A caliper of 1.5 years was applied to ensure close age matching. The matched dataset included equal numbers of postmenopausal and premenopausal women (n = 97,076 per group), and subsequent analyses of the CHIP–stroke association were performed within this matched population, further adjusting for baseline age and other covariates to minimize any residual confounding.

### Genetically predicted cytokine modifier analysis

The genetically predicted cytokines were estimated among participants of European ancestry in the BioVU (N = 107,479), AoU (N= 118,724) and UKB (N= 458,681). To derive genetically predicted cytokines, we first identified representative SNPs for each susceptibility locus based on a Bayesian ridge model trained with SomaLogic protein measurements from the INTERVAL study (OPGS000019).^38^ The genetically predicted cytokines were then calculated by summing the weighted genotypes of all selected variants using the following formula:

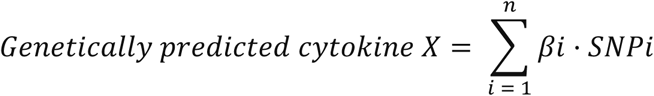

Where *βi* represents the estimated weight (i.e., the natural logarithm of the odds ratio [OR]) of the i-th SNP, derived from the reference datasets, and *SNPi* is the genotype dose of each risk allele for that SNP. In total, 27 distinct genetically predicted cytokines were computed using this approach.

For genetically predicted cytokines, analyses were performed using logistic regression testing the association between genetically predicted cytokine level and whether the participant had a stroke event, adjusting for covariates listed above. The analysis was performed separately for CHIP carriers and non-CHIP carriers. We tested statistical interaction between CHIP status and genetically predicted cytokines on stroke risk by including an interaction term in the regression model. The odds of stroke per 1 standard deviation change in cytokine levels were estimated separately in CHIP carriers and non-carriers, and interaction effects were assessed using the ratio of odds ratios and corresponding 95% confidence intervals. Multiple testing correction was applied with a Bonferroni threshold of *P* < 0.05/27. All analyses were conducted by R version 4.2.0 (https://www.r-project.org).

## Results

After applying the exclusion criteria, a total of 800,160 participants were included across three cohorts: 131,862 from BioVU, 186,910 from AoU, and 481,388 from the UKB. The mean age at blood draw was 59.2 ± 11.4 years in BioVU, 60.7 ± 11.3 years in AoU, and 56.5 ± 8.1 years in the UKB. Female accounted for 54 to 60% of participants across cohorts. Most participants were of European ancestry, although the AoU and BioVU cohorts included relatively high proportions of African ancestry individuals (19.3% and 10.4%, respectively). The median follow-up time was approximately 4.4 years (IQR 1.8–10.2) in BioVU, 1.9 years (IQR 0.4–3.9) in AoU and 12.4 years (IQR 11.7–13.1) in the UKB. During follow-up, incident stroke occurred in 7.3% of BioVU (n = 9,351), 2.7% of AoU (n = 5,054) and 0.8% of the UKB (n = 3,862) **(Table 1)**. As BioVU is derived from electronic health records of a tertiary care health system, it had the highest incidence of stroke.

**Table 1.** Basic socio-demographic characteristics by cohort.

|  | **BioVU** |  | **All of Us** |  | **UK Biobank** |  |
| --- | --- | --- | --- | --- | --- | --- |
|  | *Total*<br>*No. 131862* | *CHIP carriers*<br>*No. 9951 (7.5%)* | *Total*<br>*No. 186910* | *CHIP carriers*<br>*No. 9760 (5.2%)* | *Total*<br>*No. 481388* | *CHIP carriers*<br>*No. 15559 (3.2%)* |
| *Demographic* |  |  |  |  |  |  |
| **Age of blood draw** | 59.2 ± 11.4 | 66.5 ± 11.7 | 60.7 ± 11.3 | 67.8 ± 10.6 | 56.5 ± 8.1 | 60.7 ± 6.7 |
| **Sex** |  |  |  |  |  |  |
| *Female* | 69432 (54.0%) | 5199 (52.2%) | 112117 (60.0%) | 5470 (56.0%) | 260771 (54.2%) | 8232 (52.9%) |
| *Male* | 59205 (46.0%) | 4752 (47.8%) | 74793 (40.0%) | 4290 (44.0%) | 220617 (45.8%) | 7327 (47.1%) |
| **Ancestry** |  |  |  |  |  |  |
| *European* | 107479 (83.6%) | 8544 (85.9%) | 118724 (63.5%) | 7099 (72.7%) | 458681 (95.3%) | 15033 (96.6%) |
| *African* | 13374 (10.4%) | 879 (8.8%) | 36057 (19.3%) | 1569 (16.1%) | 9215 (1.9%) | 204 (1.3%) |
| *East Asian* | 1682 (1.3%) | 97 (1.0%) | 2853 (1.5%) | 112 (1.1%) | 13492 (2.8%) | 322 (2.1%) |
| **Smoking status** |  |  |  |  |  |  |
| *No* | 46438 (36.1%) | 2742 (37.6%) | 102413 (54.8%) | 4964 (50.9%) | 262613 (54.6%) | 7397 (47.7%) |
| *Yes* | 82199 (63.9%) | 6209 (62.4%) | 80471 (43.1%) | 4594 (47.1%) | 216317 (45.0%) | 8060 (51.9%) |
| *Baseline health status* |  |  |  |  |  |  |
| **Hypertension** | 19223 (14.9%) | 1755 (17.6%) | 8861 (4.7%) | 592 (6.1%) | 48536 (10.1%) | 2160 (13.9%) |
| **Type 2 Diabetes** | 26745 (20.8%) | 2264 (22.8%) | 25102 (13.4%) | 1330 (13.6%) | 14203 (3.0%) | 604 (3.9%) |
| **LDL-C (mmol/L)** | 2.7 ± 0.6 | 2.6 ± 0.6 | 2.7 ± 0.6 | 2.7 ± 0.7 | 3.6 ± 0.9 | 3.5 ± 0.9 |
| **BMI** | 30.8 ± 6.9 | 30.3 ± 7.1 | 29.9 ± 7.2 | 29.3 ± 6.7 | 27.4 ± 4.8 | 27.5 ± 4.6 |
| *Health outcome* |  |  |  |  |  |  |
| **Stroke incidence** |  |  |  |  |  |  |
| *Overall* | 9351 (7.3%) | 884 (8.9%) | 5054 (2.7%) | 336 (3.4%) | 3862 (0.8%) | 194 (1.2%) |
| *Ischemic stroke* | 4215 (3.3%) | 360 (3.6%) | 3338 (1.8%) | 220 (2.3%) | 1950 (0.4%) | 99 (0.6%) |
| *Hemorrhagic stroke* | 2077 (1.6%) | 187 (1.9%) | 634 (0.3%) | 47 (0.5%) | 380 (0.1%) | 21 (0.1%) |

### Baseline clonal hematopoiesis characteristics

Across the three cohorts, a total of 35,270 (4.4%) participants had ≥1 detectable CHIP mutation(s), with rates of 7.5% in BioVU, 5.2% in AoU, and 3.2% in the UKB. CHIP carriers were, on average, older than non-carriers (*P*<.001), and the prevalence was higher for male than female (*P*<.001). The mutational landscape of CHIP was dominated by *DNMT3A*, *TET2*, and *ASXL1*, which together accounted for approximately 82% of all detected mutations (**Supplementary Fig.1**). CHIP prevalence was associated with age in each cohort. Age-related increases in CHIP prevalence were consistent across cohorts, although the absolute frequencies differed, with BioVU showing the highest prevalence at all age groups. When stratified by sex, both men and women demonstrated similar age-related trends, with only modest differences in overall prevalence after 80 years (**Supplementary Fig.2**).

### CHIP associated with increased risk of incident stroke

In our primary analyses of the three cohorts, carriers of CHIP mutations had a significantly higher risk of incident stroke compared with non-carriers (HRs ranging from 1.17 to 1.37 across cohorts) (**Supplementary Fig. 3)**. To further increase power, we additionally included our previously published analysis of the WHI in our meta-analysis.^25^ The combined results demonstrated a robust association between CHIP and stroke risk (HR= 1.20, 95% CI 1.13–1.27; *P* = 2.21 × 10⁻^10^; I^2^ = 15.3%) (**Figure 2)**. We further analyzed traditional stroke risk factors within the cohorts and found that the risk associated with CHIP was comparable to that conferred by male sex (male vs. female, HR = 1.19, 95% CI 1.15–1.23) (**Supplementary Fig. 4)**.

**Figure 2.**
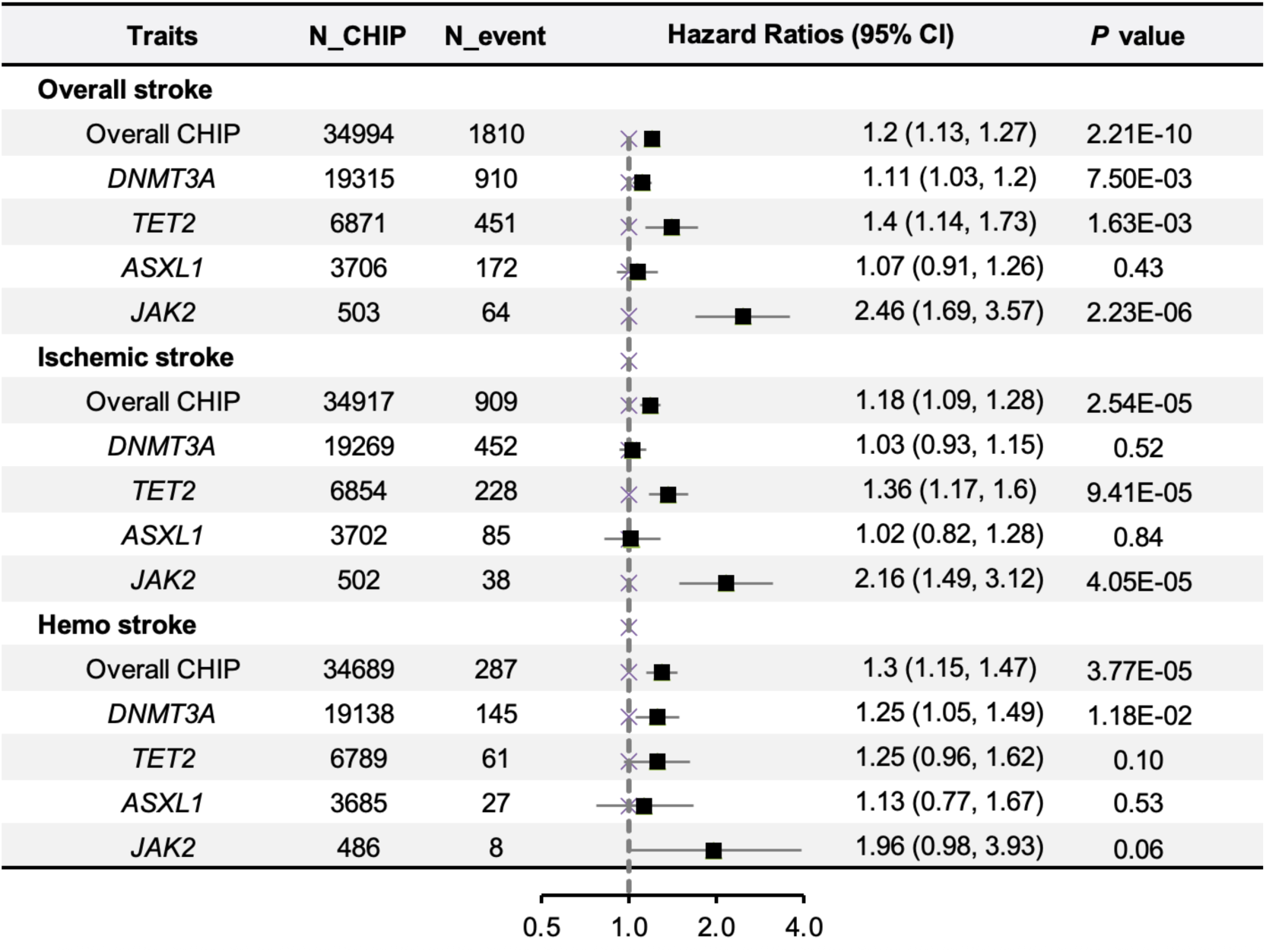
Time-to-event association of CHIP with incident stroke after meta-analysis. This forest plot shows the associations of overall CHIP and individual driver gene mutations (*DNMT3A, TET2, ASXL1, JAK2*) with stroke outcomes. The column “N_CHIP” indicates the number of CHIP carriers included, and “N_event” represents the number of stroke cases among these carriers. Hazard ratios (HRs) with 95% confidence intervals quantify the effect sizes. Results are derived from meta-analysis across the All of Us, BioVU, and UK Biobank cohorts, supplemented by published data from WHI.

Risk of stroke differed by CHIP driver mutation. *TET2* CHIP was associated with a relatively high-risk (HR= 1.40, 95% CI 1.14–1.73; *P* = 1.63× 10⁻^3^), with consistent effects across the four cohorts. *JAK2* CHIP conferred the highest increased risk (HR= 2.46, 95% CI 1.69–3.57; *P* = 2.23× 10⁻^6^), despite the relatively small number of carriers (N = 503). *DNMT3A* CHIP showed a weaker but statistically significant association (HR= 1.11, 95% CI 1.03–1.22; *P* = 7.50× 10⁻^3^), whereas *ASXL1* CHIP was not significantly associated with stroke (HR= 1.07, 95% CI 0.91–1.26, *P* = 0.43). Kaplan–Meier analyses based on the BioVU, AoU and UKB cohorts illustrate these findings, showing a lower stroke-free survival probability among CHIP carriers compared with non-carriers (log-rank *P* < .0001). The decline was particularly pronounced in individuals harboring *TET2* mutations, whose risk trajectory was substantially steeper than that of non-CHIP carriers and carriers of *DNMT3A* mutations **(Figure 3A)**.

**Figure 3.**
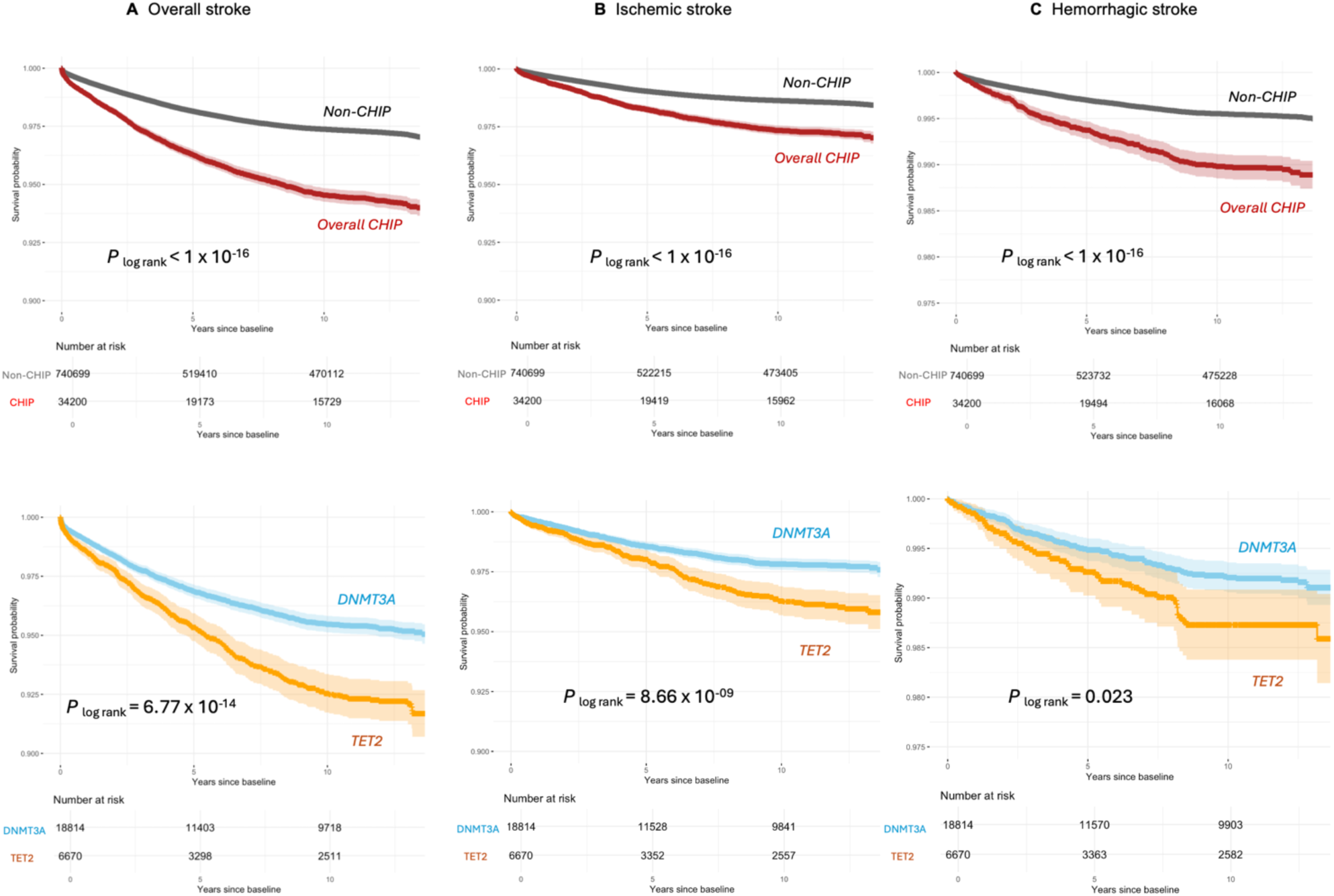
Kaplan–Meier curves for stroke risk according to CHIP status and driver genes. Kaplan–Meier analyses of (A) overall stroke, (B) ischemic stroke, and (C) hemorrhagic stroke in the combined BioVU, *All of Us*, and UK Biobank cohorts. Participants were categorized into: non-CHIP group (grey line), all CHIP carriers (red line), *DNMT3A*-CHIP carriers (blue line), and *TET2*-CHIP carriers (orange line). CHIP carriers had significantly lower stroke-free survival compared with non-carriers, with particularly steep declines among individuals harboring *TET2* mutations. The number at risk at each time point is shown below the plots.

Given that CHIP is associated with atrial fibrillation and atrial fibrillation is associated with stroke, we next performed a sensitivity analysis further adjusting for atrial fibrillation, which is not traditionally adjusted for incident stroke analyses. The association between CHIP and stroke remained significant after this additional adjustment (HR = 1.19, 95% CI 1.08–1.31, *P* = 4.81 × 10^−4^) (**Supplementary Table.3)**, indicating that the observed relationship was not driven by comorbid AF among CHIP carriers. We further examined whether the association between CHIP and stroke risk differed by clone size. Interestingly, the results were consistent across thresholds: the hazard of CHIP-associated incident stroke remained essentially unchanged when restricting to clones with VAF ≥10% (HR= 1.22, 95% CI 1.14–1.30) compared with the standard VAF ≥2% definition (HR= 1.22, 95% CI 1.12–1.32). Similar patterns were observed for individual driver genes, indicating that the increased stroke risk associated with CHIP was not materially modified by clone size **(Supplementary Fig.5)**.

### Different CHIP driver mutations associate with ischemic and hemorrhagic stroke

To determine whether the impact of CHIP varies across stroke subtypes, we analyzed its associations with ischemic and hemorrhagic stroke separately. When stratified by stroke subtype, CHIP was associated with increased risk of both ischemic and hemorrhagic stroke. The relative risk was slightly higher for hemorrhagic stroke (HR= 1.30, 95% CI 1.15–1.47; *P* = 3.77 × 10⁻^5^) **(Figure 2, Supplementary Fig.6)** than for ischemic stroke (HR= 1.18, 95% CI 1.09–1.28; *P* = 2.54 × 10⁻^5^) **(Figure 2, Supplementary Fig.7)**. In comparative analyses with established risk factors, the effect of CHIP on ischemic stroke risk was of similar magnitude to that observed for male sex (HR = 1.20), while for hemorrhagic stroke the impact of CHIP approached that of smoking status (HR = 1.28) (**Supplementary Fig. 4)**.

As with overall stroke, different drivers exhibited distinct associations. *TET2* CHIP was strongly associated with ischemic stroke (HR= 1.36, 95% CI 1.17–1.61; *P* = 9.41 × 10⁻^5^) but showed no significant association with hemorrhagic stroke (HR= 1.25, 95% CI 0.96–1.62; *P* = 0.10). *DNMT3A* CHIP was not associated with ischemic stroke (HR= 1.03, 95% CI 0.93–1.15) but demonstrated a modest association with hemorrhagic stroke (HR= 1.25, 95% CI 1.05–1.49; *P* = 0.02). *ASXL1* CHIP was not significantly related to either subtype. In contrast, *JAK2* CHIP conferred elevated risks for ischemic (HR= 2.16, 95% CI 1.49–3.12; *P* = 4.05 × 10⁻^5^) and hemorrhagic stroke (HR= 1.96, 95% CI 0.98–3.93; *P* = 0.06), with the latter not reaching statistical significance due to limited *JAK2* CHIP carriers.

CHIP is associated with atrial fibrillation,^39–41^ a stroke risk factor, so we performed additional adjustment for atrial fibrillation. The associations between CHIP and both ischemic and hemorrhagic stroke remained significant (**Supplementary Tables 4–5**). We further evaluated whether the association of CHIP with ischemic and hemorrhagic stroke differed by VAF. Consistent with the findings for overall stroke, the risk estimates were similar when restricting to large clones with VAF ≥10% compared with the standard VAF ≥2% definition, with no evidence of heterogeneity across subtypes and driver genes (**Supplementary Figure 8-9**).

### Effect of menopausal status on the association between CHIP and stroke

To assess sex-specific effects, we stratified the analyses by sex within each cohort. Across BioVU, AoU and UKB, CHIP carriers of both sexes had a significantly higher risk of incident stroke than non-carriers. In the meta-analysis, the association of CHIP and stroke incidence was significant in men (HR= 1.26, 95% CI 1.15– 1.39; *P* = 2.10 × 10^−6^) and in women (HR= 1.17, 95% CI 1.08–1.27; *P* = 1.11 × 10^−4^), with a numerically higher hazard in men **(Figure 4A)**. To examine whether menopausal status modifies the association between CHIP and stroke, we stratified female participants in the UKB by baseline menopausal status. Among premenopausal women, CHIP was not significantly associated with incident stroke (HR = 0.70, 95% CI 0.36–1.43, *P* = 0.33), whereas the association was significant among postmenopausal women (HR = 1.49, 95% CI 1.16–1.92, *P* = 1.91 × 10^−3^). To minimize potential confounding by age, we conducted a 1:1 nearest-neighbor matching baseline age between the two groups. The results remained consistent: CHIP was significantly associated with higher stroke risk in postmenopausal women (HR = 1.46, 95% CI 1.03–2.08, *P* = 0.033), but not in premenopausal women (*P* = 0.34) **(Figure 4B)**.

**Figure 4.**
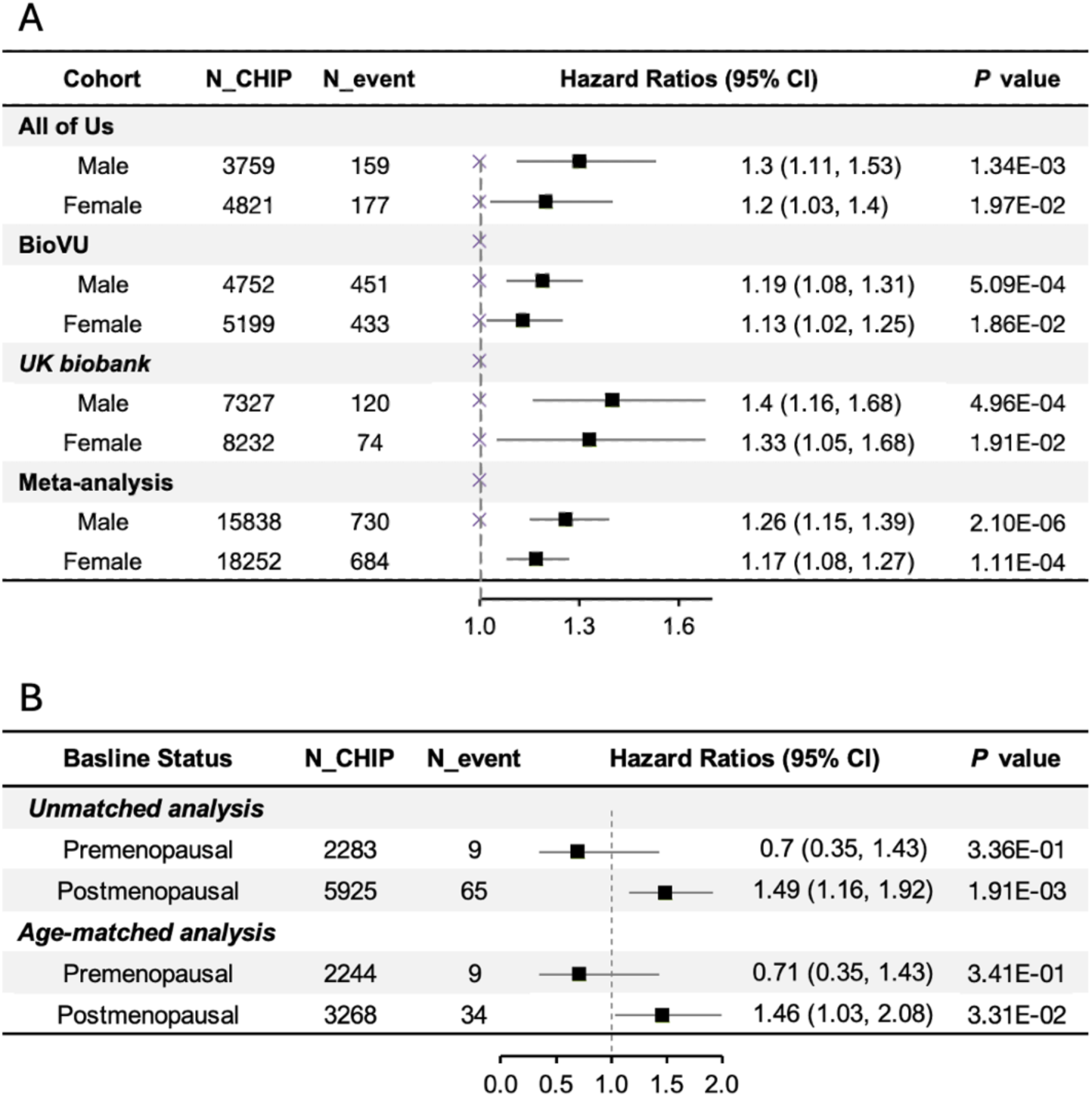
Sex-stratified associations of CHIP with incident stroke. (A) Forest plots showing hazard ratios and 95% CIs for the association of CHIP with incident stroke, stratified by sex across the BioVU, *All of Us*, and UK Biobank cohorts, and combined in meta-analysis. (B) Hazard ratios and 95% CIs for the association between CHIP and incident stroke are shown separately for premenopausal and postmenopausal women in UKB. Results are presented for both the unmatched and age-matched analyses. The same Cox regression model was applied in both analyses with the same covariates (including baseline age). The column “N_CHIP” indicates the number of CHIP carriers included, and “N_event” represents the number of stroke cases among CHIP carriers.

### Genetically predicted cytokines and stroke risk among CHIP carriers

Finally, we investigated the role of genetically predicted circulating levels of 27 cytokines in mediating the association between CHIP and stroke. Among the proteins evaluated, genetically predicted IL-1 receptor accessory protein (gIL-1RAP) was significantly associated with increased stroke risk (OR= 1.17, 95% CI 1.06– 1.28; *P* = 1.50 × 10⁻^3^) (**Figure 5**), whereas no significant associations were observed for other cytokines (**Supplementary Figure 10**). Notably, this association was restricted to CHIP carriers and was not observed in the overall population (OR = 1.03; *P* = 0.68) or in non-carriers (OR = 0.99; *P* = 0.55). We observed a significant positive interaction between CHIP status and gIL-1RAP on stroke risk, indicating an amplifying effect of gIL-1RAP among CHIP carriers (interaction HR = 1.18, 95% CI: 1.07–1.30; *P* for interaction = 6.06 × 10^−4^). A similar pattern was observed for ischemic stroke, with a significant CHIP–IL-1RAP interaction (interaction HR = 1.16, 95% CI: 1.03–1.31; *P* for interaction = 0.014). For hemorrhagic stroke, the direction of effect was consistent, but the interaction did not reach statistical significance (interaction HR = 1.25, 95% CI: 0.99–1.57; *P* for interaction = 0.066).

**Figure 5.**
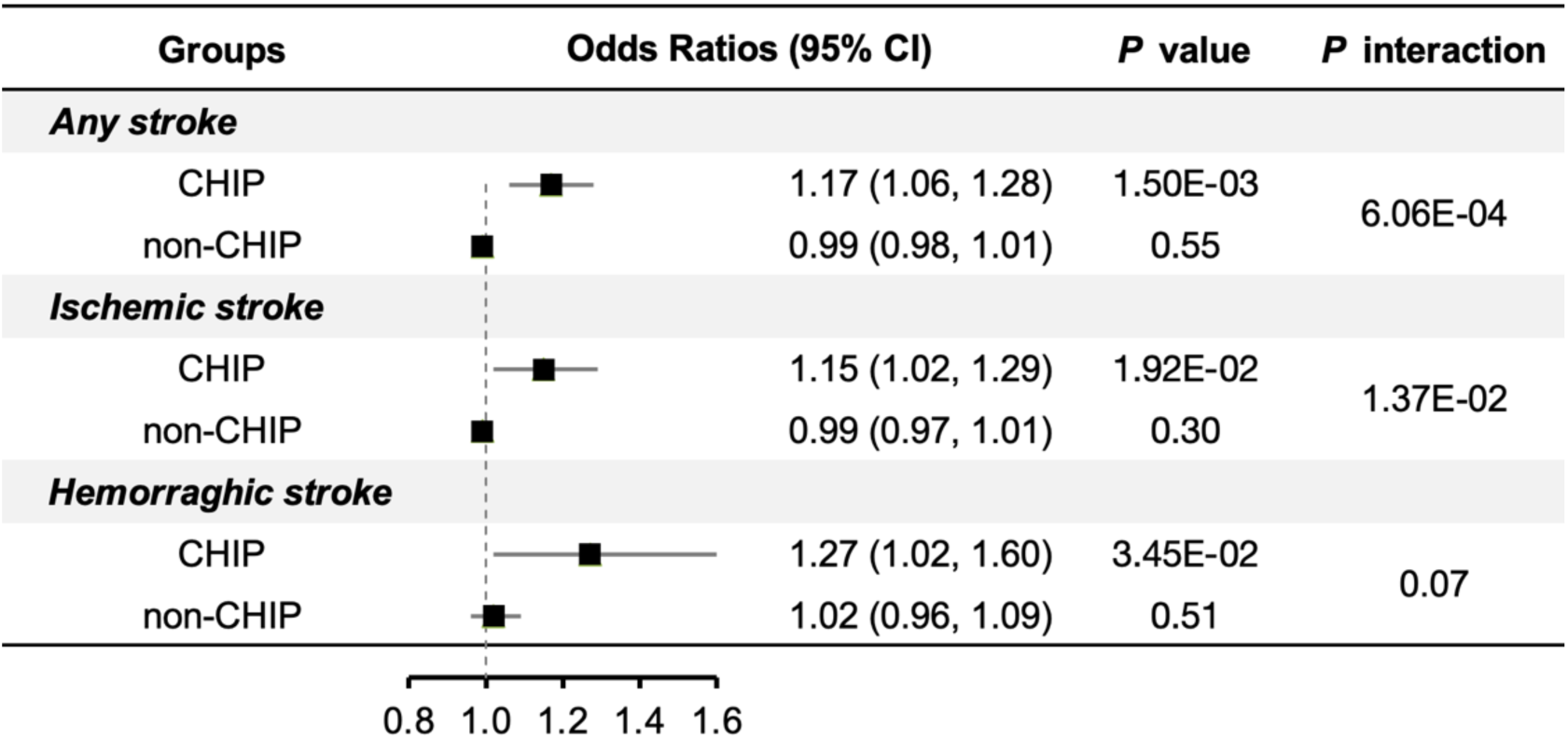
Association of genetically predicted IL-1 receptor accessory protein with stroke risk. Forest plots display the associations between genetically predicted interleukin-1 receptor accessory protein (IL-1RAP) levels and risk of stroke among CHIP carriers and non-carriers. Odds ratios and 95% CIs were estimated using logistic regression models adjusted for age, age2, sex, smoking status, BMI, LDL_C, baseline hypertension and type 2 diabetes. Results are derived from meta-analysis across the BioVU, AoU, and UKB cohorts restricted to participants of European ancestry.

## Discussion

In this large-scale analysis across three independent cohorts, we found that CHIP was significantly associated with an increased risk of incident stroke. This association was consistent across cohorts and extended to both ischemic and hemorrhagic subtypes, with relatively higher hazard ratios observed for hemorrhagic stroke. Gene-specific analyses indicated that mutations in *JAK2* and *TET2* were the strongest drivers of stroke risk, whereas *DNMT3A* exerted weaker effects. Analyses stratified by sex and then menopausal status revealed that CHIP was associated with stroke risk in both sexes; however, among women, this association was evident only in those who were postmenopausal. IL-1RAP, a key component of the interleukin-1 signaling pathway, is a potential modifier which enhances risk of CHIP-related stroke risk. These findings permit several conclusions.

First, CHIP is a strong risk factor for incident stroke, with a hazard ratio comparable to traditional stroke risk factors such as male sex for ischemic stroke and smoking for hemorrhagic stroke. Earlier reports suggested strong associations in smaller cohorts,^24^ whereas larger population-based studies yielded more modest or null effects, particularly outside of *JAK2* mutations.^32,33^ Interestingly, the hazard ratios were numerically higher for hemorrhagic stroke, which may reflect CHIP-driven inflammatory activation that compromises vascular integrity.^25,42,43^ CHIP-associated stroke risk varies substantially by driver gene and stroke type. While it is well-known that *JAK2* CHIP strongly increases risk of ischemic stroke, the relationship of other driver genes and both types of stroke are more contested. Our results suggest that *TET2* CHIP confers a substantial risk of incident ischemic stroke, but not hemorrhagic stroke. In contrast, *DNMT3A* CHIP confers a risk of incident hemorrhagic stroke, but not ischemic stroke.

Second, our subgroup analyses also revealed important insights regarding clone size and hormonal status. The associations of CHIP with stroke were stable across variant allele frequency thresholds (≥2% and ≥10%). These results suggest that rather than the proportion of mutated peripheral blood cells, the systemic inflammatory signaling present in anyone with CHIP may be contributing to stroke risk. This may reflect the long latency between CHIP acquisition and the onset of overt vascular disease, during which even small clones can chronically contribute to systemic inflammation. Although the confidence intervals of the estimates overlapped, the point estimates suggested that women may have a lower CHIP-attributable risk of stroke than men. When we further hypothesized that estrogen may play a protective role against CHIP-associated stroke risk, we found that in women, the association between CHIP and stroke was evident only among those who were postmenopausal, whereas no significant association was observed in premenopausal women. We observed the same findings after matching for age. However, disentangling the association of two age-associated phenomena in CHIP and menopause remains challenging. While these results are far from definitive, they suggest that further work should be undertaken to understand how loss of endogenous estrogen, a hormone with known anti-inflammatory, antioxidative, and vasculoprotective effects^44–46^, may amplify the vascular consequences of CHIP-related inflammation.

Third, IL-1 signaling may be a promising drug target for CHIP-associated stroke risk with biologics such as anakinra or canakinumab. CHIP carriers with higher genetically predicted IL1-RAP signaling experience heightened stroke risk. IL-1RAP is an essential co-receptor for IL-1 family cytokines, including IL-1, IL-33, and IL-36, and thus amplifies interleukin-driven immune responses.^47^ Our human genetic evidence is corroborated by prior work examining usage of IL-1 blockade in the Canakinumab Anti-inflammatory Thrombosis Outcomes (CANTOS) trial to reduce CHIP-associated disease risk. In those with CHIP, administration of canakinumab demonstrated reductions in major adverse cardiovascular events,^48^ inflammatory signaling,^49^ and incident solid-organ malignancies^50^ exclusively among CHIP carriers. Future work is needed to test prospectively or via analysis of prior samples whether IL-1 blockade may have similar effects for incident stroke.

### Strengths and limitations

Our study has several notable strengths. We analyzed an exceptionally large sample of more than 800,000 participants, which provided statistical power to detect modest associations and examine rare CHIP mutations. CHIP identification and stroke outcomes were harmonized across cohorts using standardized sequencing pipelines and definitions. We further adjusted for an extensive set of vascular and demographic covariates and performed detailed gene-specific, variant allele frequency, and sex-stratified analyses. Nevertheless, important limitations should be acknowledged. First, some controls may have CHIP mutations with low VAF, which cause us to underestimate CHIP-associated stroke risk, as CHIP was detected from whole-genome and whole-exome sequencing with standard depth, rather than the gold-standard deep sequencing.^51^ Second, only a single baseline sequencing measurement was available, precluding assessment of clonal dynamics over time. Third, cytokine levels were genetically predicted rather than directly measured and thus may not fully capture circulating concentrations or context-specific regulation. Fourth, only 9 stroke events occurred in pre-menopausal CHIP women, as CHIP was relatively uncommon among premenopausal individuals. These caveats underscore the need for longitudinal sequencing, direct biomarker profiling, and experimental validation to refine our understanding of the mechanisms linking CHIP to stroke.

## Conclusion

In conclusion, this study establishes clonal hematopoiesis as an important determinant of incident stroke risk in a genotype-specific manner, with comparable effect size as traditional risk factors. Our findings suggest that future work should investigate the synergistic effects of CHIP and menopause as women age on stroke and cardiovascular risk. Finally, our human genetic evidence nominates IL-1 as a therapeutic target for reduction in CHIP-associated stroke risk. These findings underscore the broader relevance of somatic mutations in stroke risk and point toward new opportunities for preventing stroke through genomically informed strategies.

## Data Availability

Individual-level sequence data and CHIP calls have been deposited with UK Biobank and are available to approved researchers by application (https://www.ukbiobank.ac.uk/register-apply/). Vanderbilt BioVU data are available through an application to the Vanderbilt Institute for Clinical and Translational Research (VICTR) BioVU Review Committee.

## Supplementary Figures

**Supplementary Fig 1.**
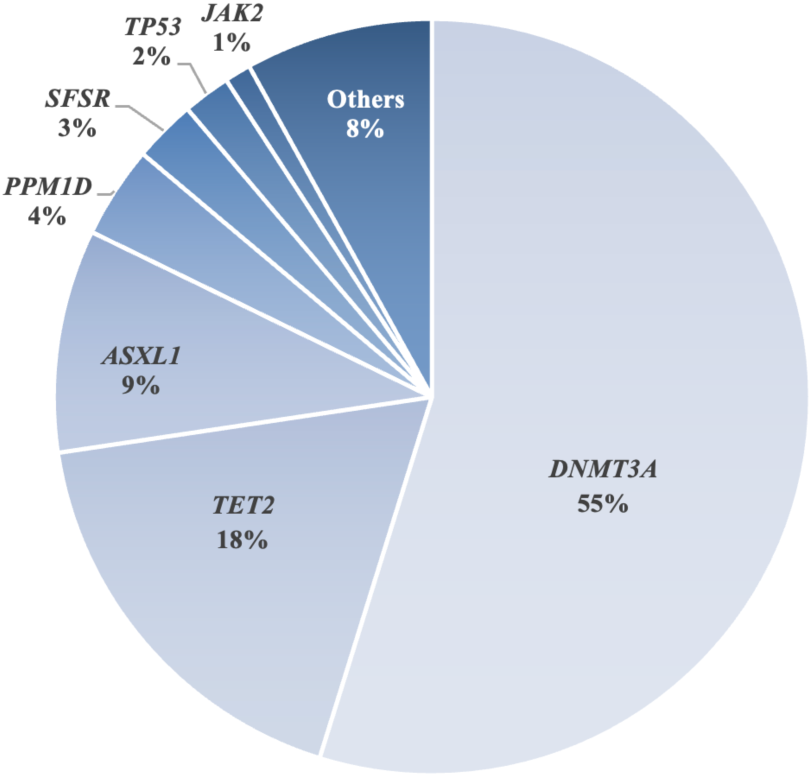
Distribution of driver gene mutations in CHIP carriers. Pie chart showing the relative frequencies of driver gene mutations among CHIP carriers. The distribution was dominated by *DNMT3A* (55%), *TET2* (18%), and *ASXL1* (9%), which together accounted for ∼82% of all detected mutations. Less frequent mutations included *PPM1D* (4%), *SRSF2* (3%), *TP53* (2%), *JAK2* (1%), and others (8%).

**Supplementary Figure 2.**
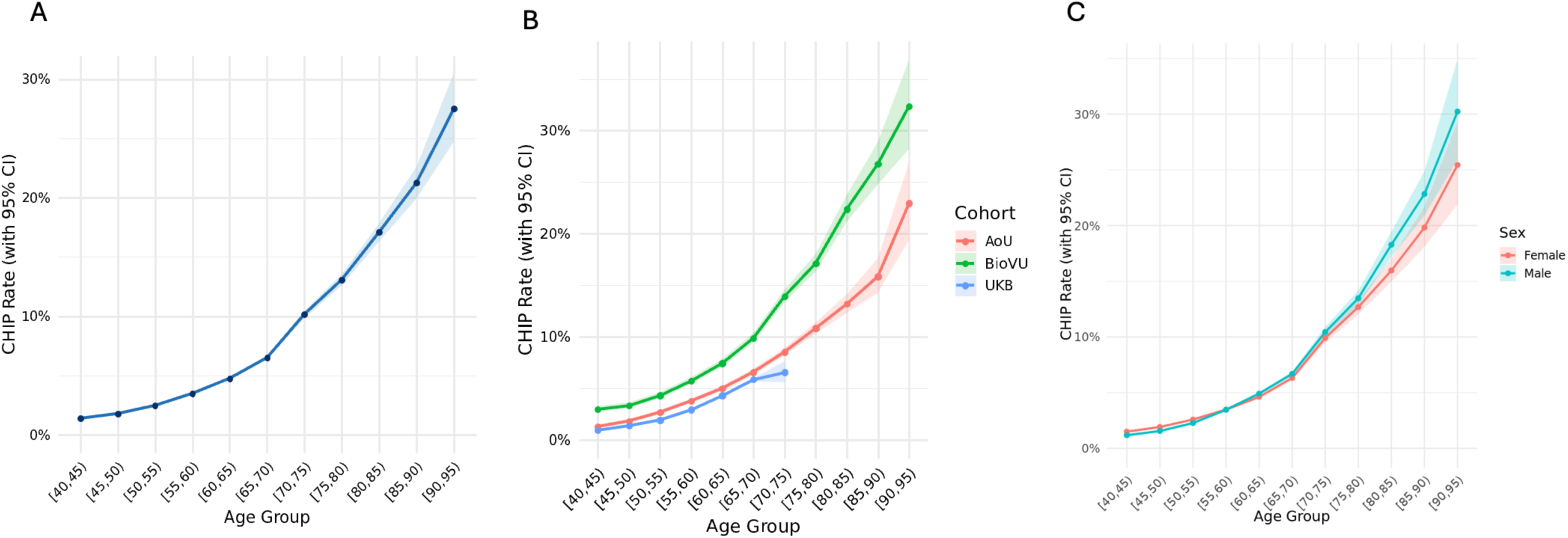
Age-related prevalence of CHIP across cohorts and by sex. (A) Overall prevalence of CHIP increased steadily with advancing age, exceeding 10% after age 70. (B) Age-stratified CHIP prevalence across the BioVU, *All of Us*, and UK Biobank cohorts, showing consistent age-dependent increases but higher absolute rates in BioVU. (C) Age-stratified CHIP prevalence by sex, demonstrating similar age-related patterns in both men and women, with slightly higher rates observed among men at older ages. Shaded areas represent 95% confidence intervals.

**Supplementary Figure 3.**
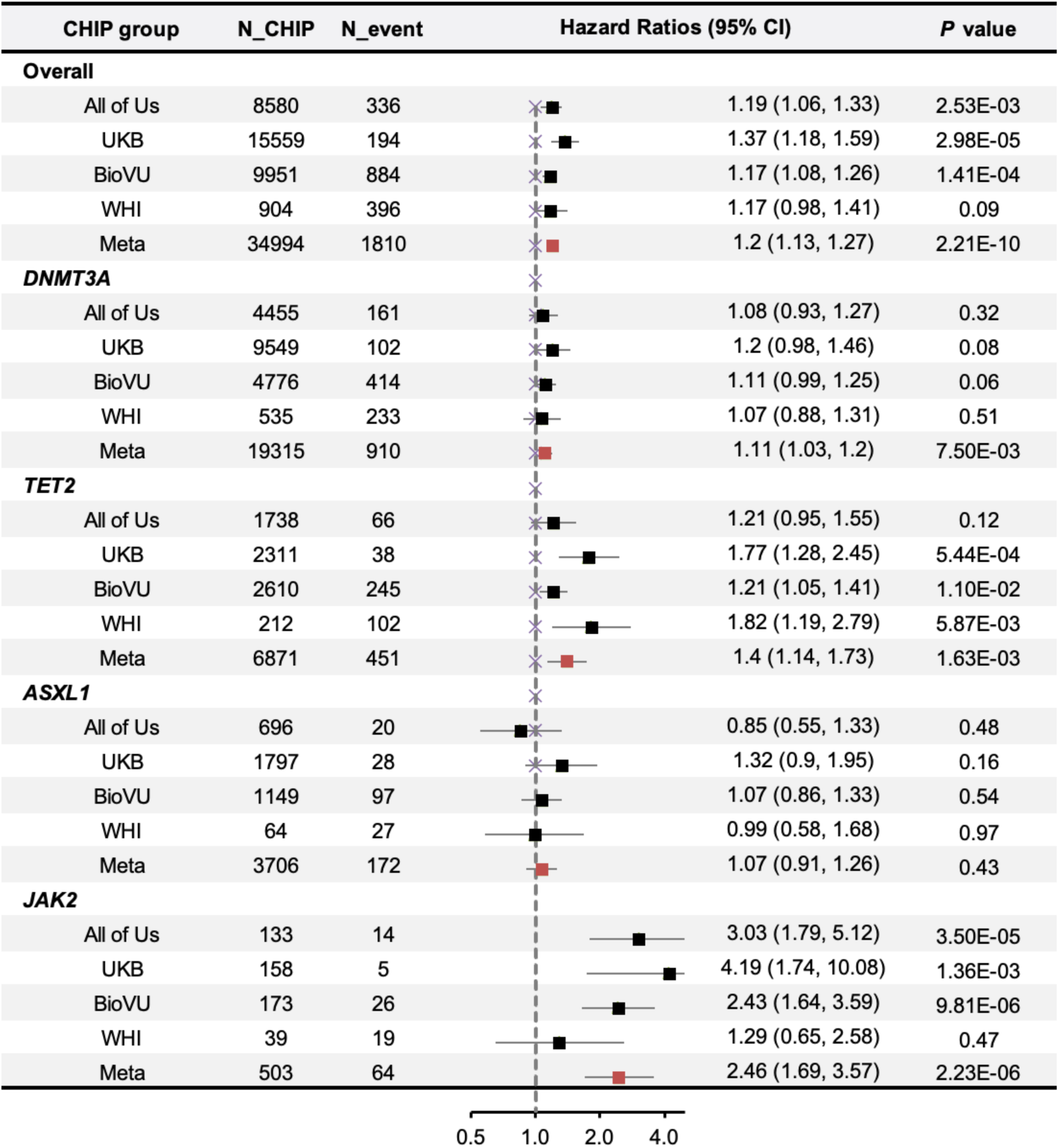
Time-to-event association of CHIP with incident overall stroke. Forest plots showing HRs and 95% CIs for the association of CHIP overall and by major driver mutations (*DNMT3A, TET2, ASXL1, JAK2*) with incident hemorrhagic stroke across the *All of Us*, UK Biobank, BioVU, and WHI cohorts, followed by meta-analysis estimates. The column “N_CHIP” indicates the number of CHIP carriers included, and “N_event” represents the number of stroke cases among these carriers. Hazard ratios (HRs) with 95% confidence intervals quantify the effect sizes.

**Supplementary Figure 4.**
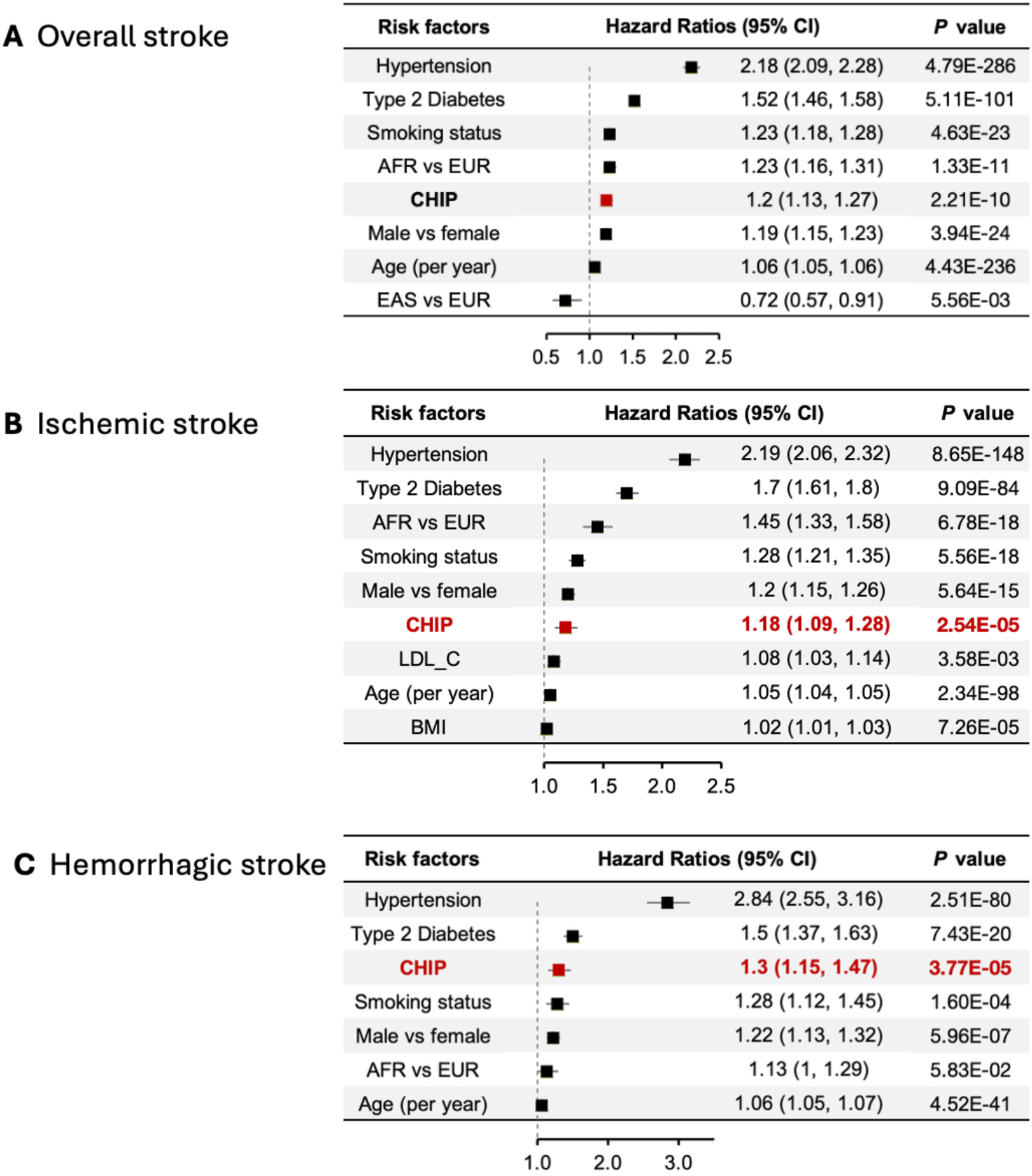
Comparative associations of CHIP and traditional vascular risk factors with stroke incidence. Forest plots show hazard ratios for the associations of CHIP (highlighted in red) and established vascular risk factors with (A) overall stroke, (B) ischemic stroke, and (C) hemorrhagic stroke. Results are based on meta-analyses of the All of Us, UK Biobank, and BioVU cohorts.

**Supplementary Figure 5.**
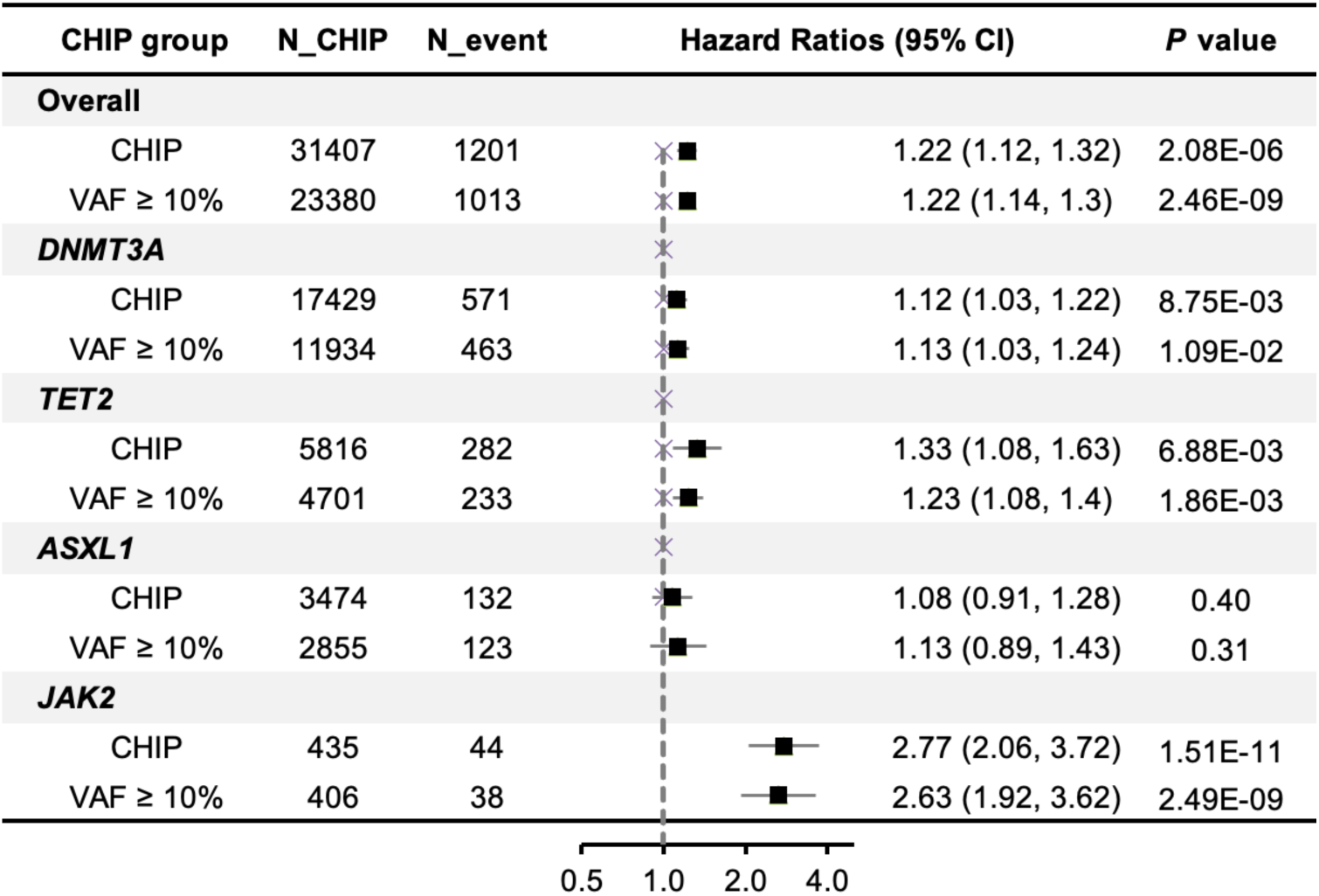
Association of CHIP with incident stroke stratified by clonal size. Forest plots showing hazard ratios and 95% CIs for the association between CHIP and incident stroke overall and by major driver genes, stratified by variant allele frequency (VAF ≥2% vs VAF ≥10%). The associations of CHIP with stroke risk were consistent across clone size thresholds, suggesting that even relatively small clones confer elevated risk.

**Supplementary Figure 6.**
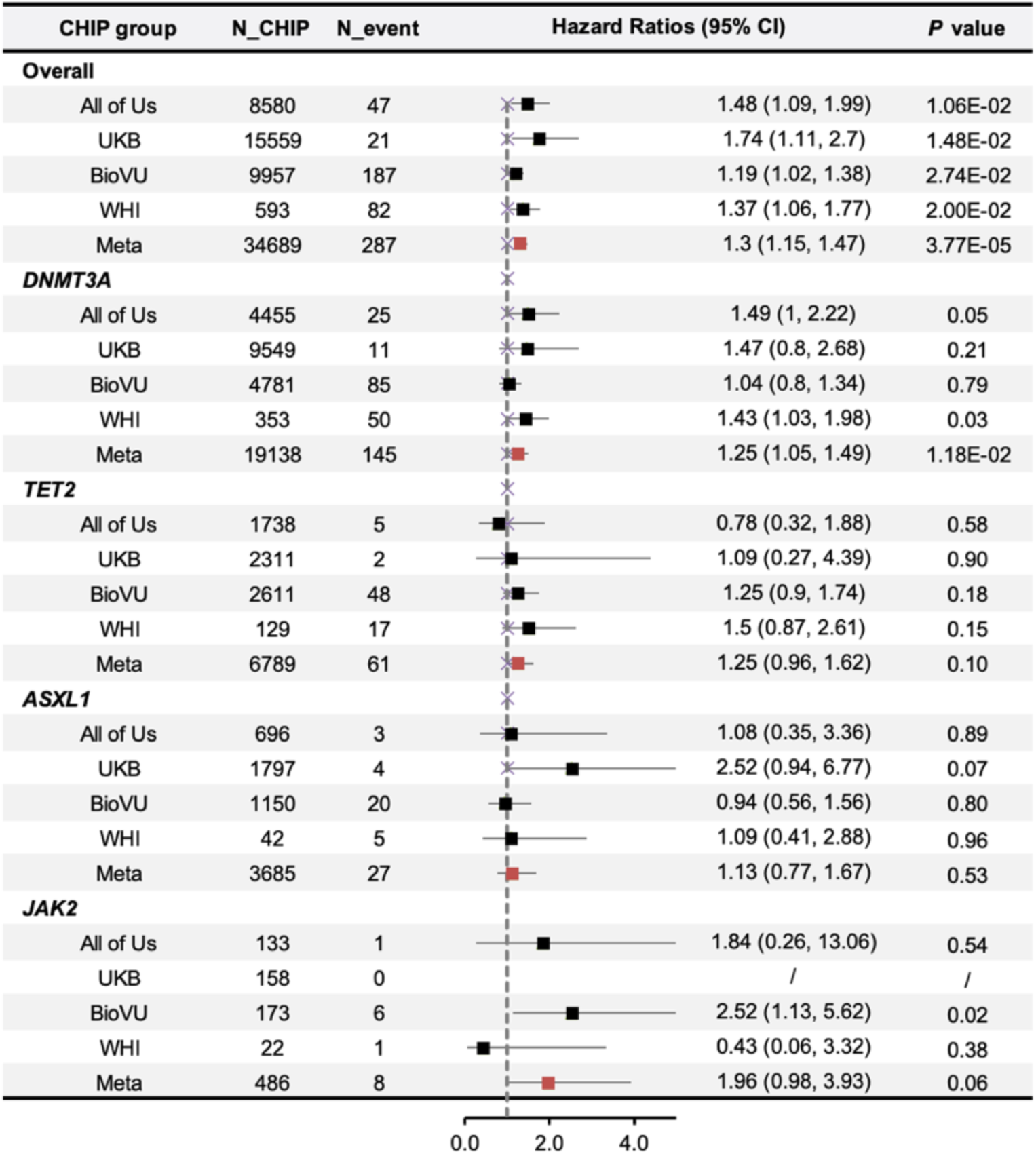
Time-to-event association of CHIP with incident hemorrhagic stroke. Forest plots showing HRs and 95% CIs for the association of CHIP overall and by major driver mutations (*DNMT3A, TET2, ASXL1, JAK2*) with incident hemorrhagic stroke across the *All of Us*, UK Biobank, BioVU, and WHI cohorts, followed by meta-analysis estimates. The column “N_CHIP” indicates the number of CHIP carriers included, and “N_event” represents the number of stroke cases among these carriers. Hazard ratios (HRs) with 95% confidence intervals quantify the effect sizes.

**Supplementary Figure 7.**
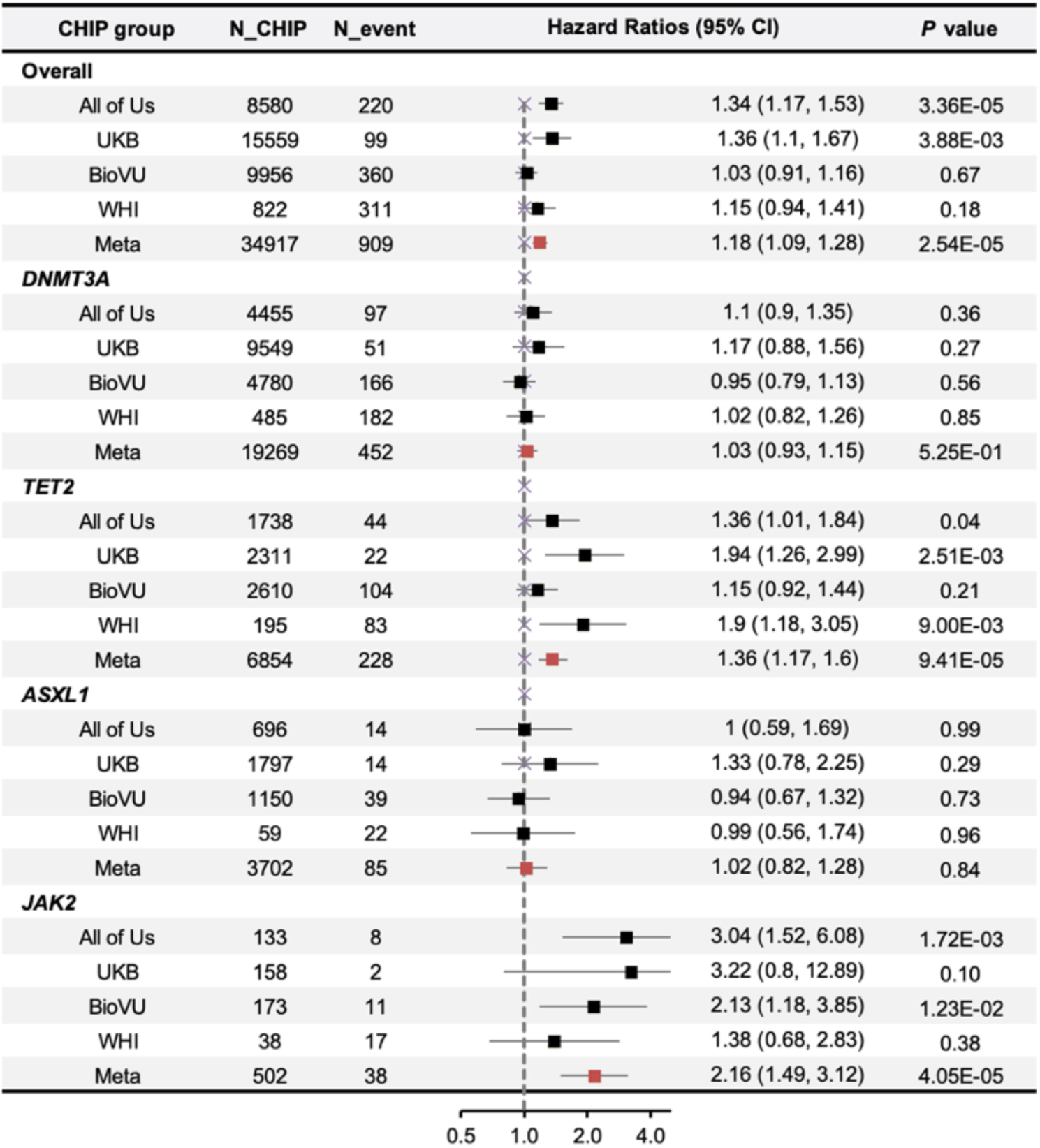
Time-to-event association of CHIP with incident ischemic stroke. Forest plots showing hazard ratios and 95%CIs for the association of CHIP overall and by major driver mutations (*DNMT3A, TET2, ASXL1, JAK2*) with incident ischemic stroke across the All of Us, UK Biobank, BioVU, and WHI cohorts, followed by meta-analysis estimates. The column “N_CHIP” indicates the number of CHIP carriers included, and “N_event” represents the number of stroke cases among these carriers. Hazard ratios (HRs) with 95% confidence intervals quantify the effect sizes.

**Supplementary Figure 8.**
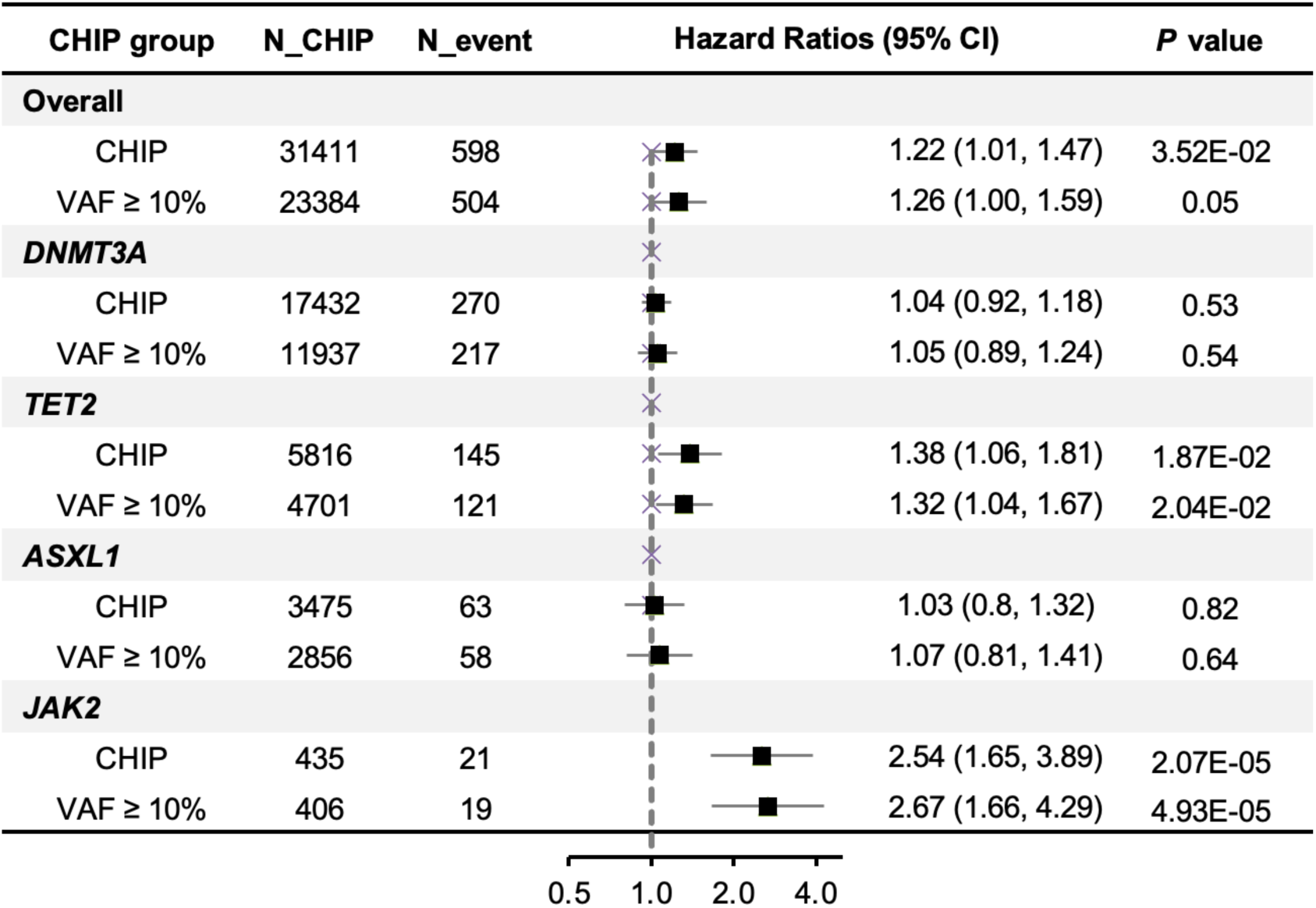
Association of CHIP with incident ischemic stroke stratified by clonal size. Forest plots showing hazard ratios and 95% CIs for the association between CHIP and incident ischemic stroke overall and by major driver genes, stratified by variant allele frequency (VAF ≥2% vs VAF ≥10%). The associations of CHIP with ischemic stroke risk were consistent across clone size thresholds, suggesting that even relatively small clones confer elevated risk.

**Supplementary Figure 9.**
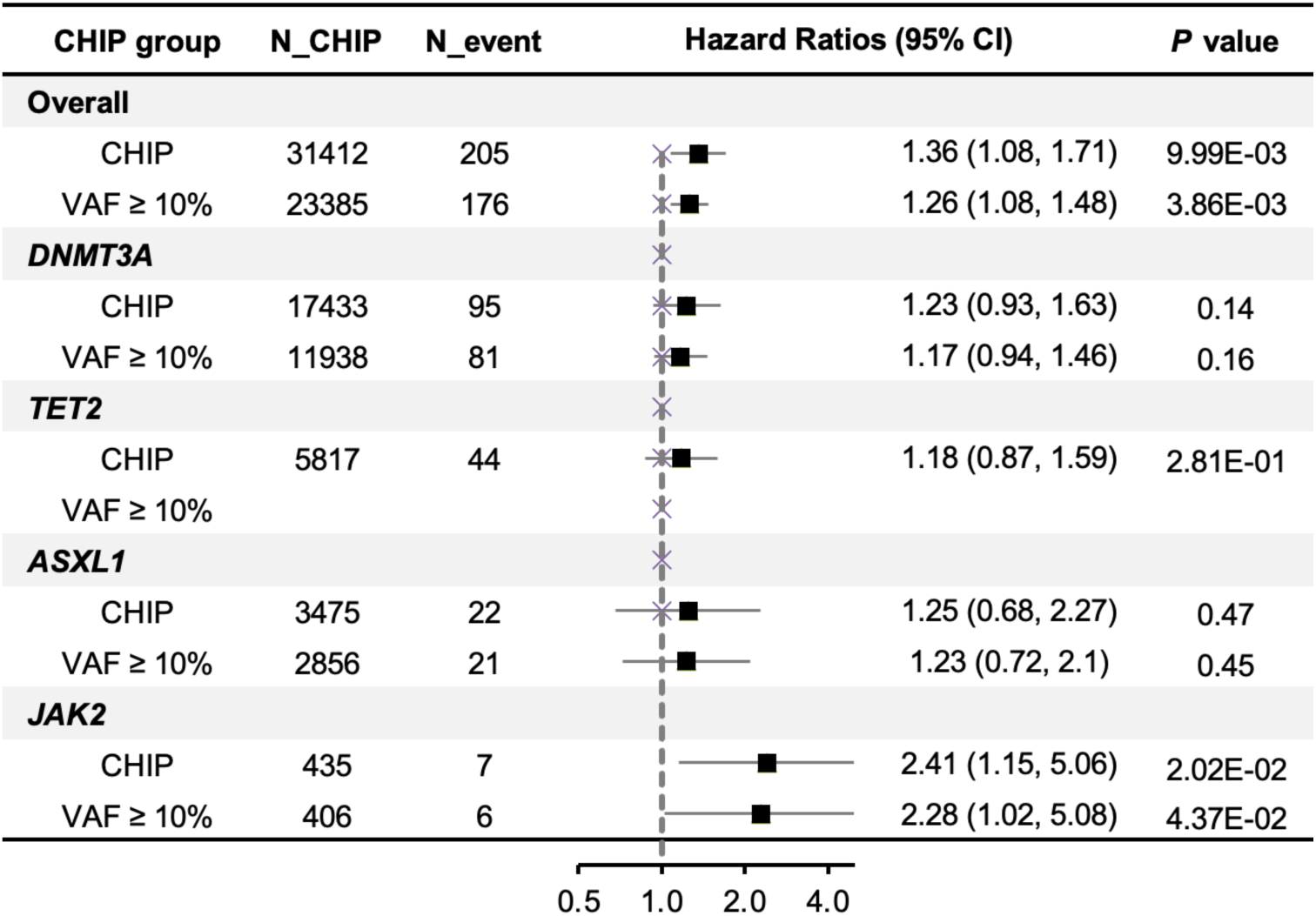
Association of CHIP with incident hemorrhagic stroke stratified by clonal size. Forest plots showing hazard ratios and 95% CIs for the association between CHIP and incident hemorrhagic stroke overall and by major driver genes, stratified by variant allele frequency (VAF ≥2% vs VAF ≥10%). The associations of CHIP with hemorrhagic stroke risk were consistent across clone size thresholds, suggesting that even relatively small clones confer elevated risks.

**Supplementary Figure 10.**
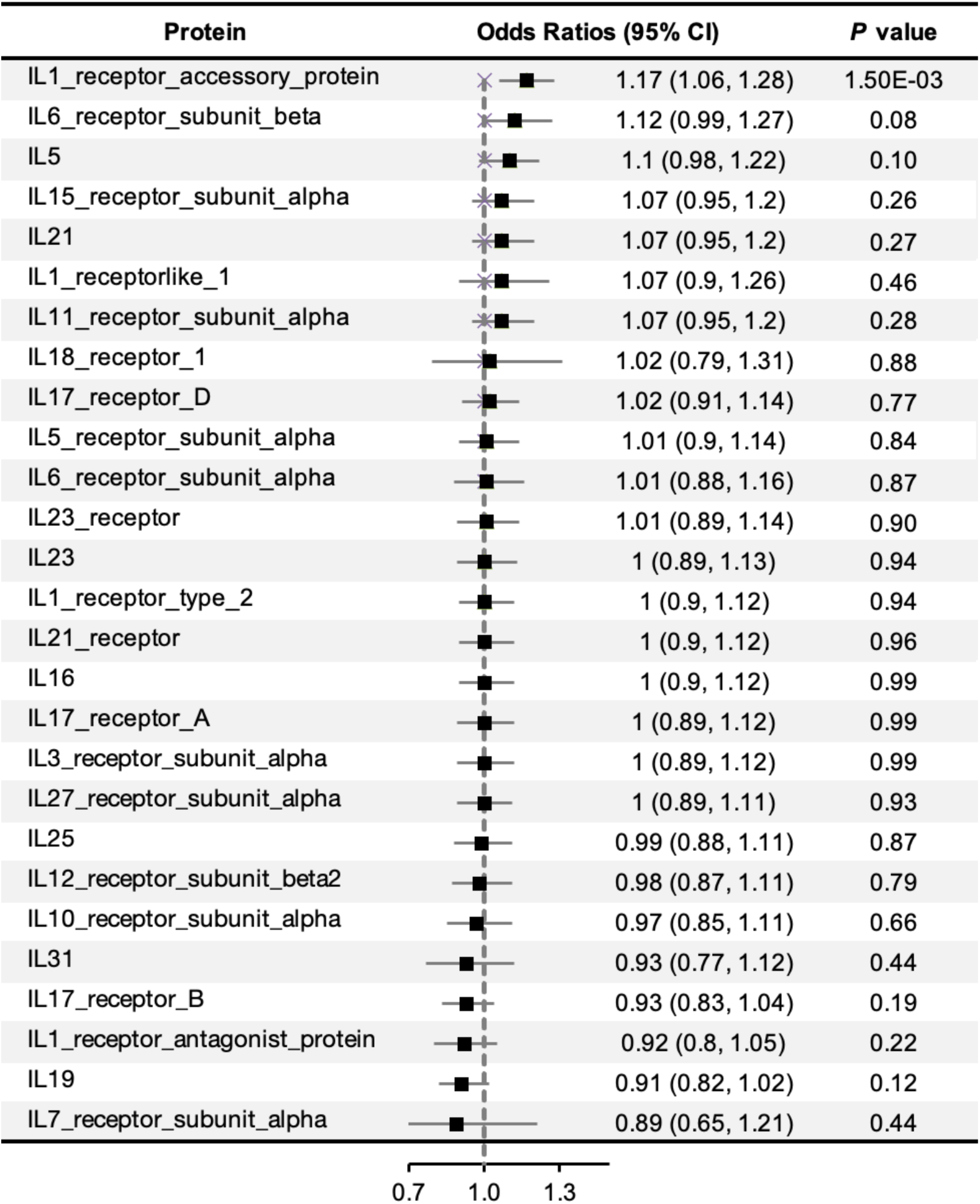
Association of genetically predicted cytokines level with stroke risk among CHIP carriers. Forest plots showing odds ratios and 95% CIs for the association between genetically predicted levels of 27 cytokines and stroke risk in CHIP carriers. Among these, only IL-1 receptor accessory protein (IL-1RAP) was significantly associated with increased risk of stroke after multiple testing correction, implicating IL-1–mediated inflammation as a potential mechanistic pathway.

## Reference

1. Feigin VL, Brainin M, Norrving B, et al. World Stroke Organization: Global Stroke Fact Sheet 2025. International Journal of Stroke. 2025;20:132–144.

2. Feigin VL, Abate MD, Abate YH, et al. Global, regional, and national burden of stroke and its risk factors, 1990–2021: a systematic analysis for the Global Burden of Disease Study 2021. The Lancet Neurology. 2024;23:973–1003.

3. Hilkens NA, Casolla B, Leung TW, De Leeuw F-E. Stroke. The Lancet. 2024;403:2820–2836.

4. Rexrode KM, Madsen TE, Yu AYX, Carcel C, Lichtman JH, Miller EC. The Impact of Sex and Gender on Stroke. Circulation Research. 2022;130:512–528.

5. Ananth CV, Brandt JS, Keyes KM, Graham HL, Kostis JB, Kostis WJ. Epidemiology and trends in stroke mortality in the USA, 1975–2019. International Journal of Epidemiology. 2023;52:858–866.

6. Eriksson M, Åsberg S, Sunnerhagen KS, Von Euler M, on behalf of the Riksstroke Collaboration. Sex Differences in Stroke Care and Outcome 2005–2018: Observations From the Swedish Stroke Register. Stroke. 2021;52:3233–3242.

7. Virani SS, Alonso A, Aparicio HJ, et al. Heart Disease and Stroke Statistics—2021 Update: A Report From the American Heart Association. Circulation. 2021;143.

8. Elkind MSV. Inflammatory Mechanisms of Stroke. Stroke. 2010;41.

9. Lindsberg PJ, Grau AJ. Inflammation and Infections as Risk Factors for Ischemic Stroke. Stroke. 2003;34:2518–2532.

10. Zhu H, Hu S, Li Y, et al. Interleukins and Ischemic Stroke. Front Immunol. 2022;13:828447.

11. Zhang L, Omarov M, Xu L, deGoma E, Natarajan P, Georgakis MK. IL6 genetic perturbation mimicking IL-6 inhibition is associated with lower cardiometabolic risk. Nat Cardiovasc Res. 2025;4:1172–1186.

12. Bick AG, Pirruccello JP, Griffin GK, et al. Genetic Interleukin 6 Signaling Deficiency Attenuates Cardiovascular Risk in Clonal Hematopoiesis. Circulation. 2020;141:124–131.

13. Kazmi S, Salehi-Pourmehr H, Sadigh-Eteghad S, Farhoudi M. The efficacy and safety of interleukin-1 receptor antagonist in stroke patients: A systematic review. Journal of Clinical Neuroscience. 2024;120:120–128.

14. Sobowale OA, Parry-Jones AR, Smith CJ, Tyrrell PJ, Rothwell NJ, Allan SM. Interleukin-1 in Stroke: From Bench to Bedside. Stroke. 2016;47:2160–2167.

15. Matys P, Mirończuk A, Starosz A, et al. Expanding Role of Interleukin-1 Family Cytokines in Acute Ischemic Stroke. IJMS. 2024;25:10515.

16. Papadopoulos A, Palaiopanos K, Björkbacka H, et al. Circulating Interleukin-6 Levels and Incident Ischemic Stroke: A Systematic Review and Meta-analysis of Prospective Studies. Neurology. 2022;98.

17. Georgakis MK, Malik R, Gill D, et al. Interleukin-6 Signaling Effects on Ischemic Stroke and Other Cardiovascular Outcomes: A Mendelian Randomization Study. Circ: Genomic and Precision Medicine. 2020;13:e002872.

18. Pawluk H, Woźniak A, Tafelska-Kaczmarek A, et al. The Role of IL-6 in Ischemic Stroke. Biomolecules. 2025;15:470.

19. Sun W, Wang S, Nan S. The Prognostic Determinant of Interleukin-10 in Patients with Acute Ischemic Stroke: An Analysis from the Perspective of Disease Management Tu W-J, editor. Disease Markers. 2021;2021:1–9.

20. Piepke M, Clausen BH, Ludewig P, et al. Interleukin-10 improves stroke outcome by controlling the detrimental Interleukin-17A response. J Neuroinflammation. 2021;18:265.

21. Mishra A, Malik R, Hachiya T, et al. Stroke genetics informs drug discovery and risk prediction across ancestries. Nature. 2022;611:115–123.

22. Malik R, Chauhan G, Traylor M, et al. Multiancestry genome-wide association study of 520,000 subjects identifies 32 loci associated with stroke and stroke subtypes. Nat Genet. 2018;50:524–537.

23. Debette S, Paré G. Stroke Genetics, Genomics, and Precision Medicine. Stroke. 2024;55:2163–2168.

24. Jaiswal S, Fontanillas P, Flannick J, et al. Age-Related Clonal Hematopoiesis Associated with Adverse Outcomes. N Engl J Med. 2014;371:2488–2498.

25. Bhattacharya R, Zekavat SM, Haessler J, et al. Clonal Hematopoiesis Is Associated With Higher Risk of Stroke. Stroke. 2022;53:788–797.

26. Jaiswal S, Ebert BL. Clonal hematopoiesis in human aging and disease. Science. 2019;366:eaan4673.

27. Bick AG, Weinstock JS, Nandakumar SK, et al. Inherited causes of clonal haematopoiesis in 97,691 whole genomes. Nature. 2020;586:763–768.

28. Jaiswal S, Natarajan P, Silver AJ, et al. Clonal Hematopoiesis and Risk of Atherosclerotic Cardiovascular Disease. N Engl J Med. 2017;377:111–121.

29. Oren O, Small AM, Libby P. Clonal hematopoiesis and atherosclerosis. Journal of Clinical Investigation. 2024;134:e180066.

30. Yu Z, Fidler TP, Ruan Y, et al. Genetic modification of inflammation- and clonal hematopoiesis-associated cardiovascular risk. J Clin Invest. 2023;133.

31. Zhao K, Shen X, Liu H, et al. Somatic and Germline Variants and Coronary Heart Disease in a Chinese Population. JAMA Cardiol. 2024;9:233.

32. Kessler MD, Damask A, O’Keeffe S, et al. Common and rare variant associations with clonal haematopoiesis phenotypes. Nature. 2022;612:301–309.

33. Kar SP, Quiros PM, Gu M, et al. Genome-wide analyses of 200,453 individuals yield new insights into the causes and consequences of clonal hematopoiesis. Nat Genet. 2022;54:1155–1166.

34. Arends CM, Liman TG, Strzelecka PM, et al. Associations of clonal hematopoiesis with recurrent vascular events and death in patients with incident ischemic stroke. Blood. 2023;141:787–799.

35. Tan H, Zhu F, Yan H, et al. Genetic Associations of Clonal Hematopoiesis With Cardioembolic Stroke: Insights From Genome-Wide Mendelian Randomization, Bulk RNA, Single-Cell RNA Sequencing. CNS Neurosci Ther. 2025;31:e70515.

36. Vlasschaert C, Mack T, Heimlich JB, et al. A practical approach to curate clonal hematopoiesis of indeterminate potential in human genetic data sets. Blood. 2023;141:2214–2223.

37. Zhao K, Hsu C-C, Pershad Y, et al. Interleukin-17 receptor-A signalling: atheroprotective role in JAK2 clonal haematopoiesis. European Heart Journal. 2025:ehaf737.

38. Xu Y, Ritchie SC, Liang Y, et al. An atlas of genetic scores to predict multi-omic traits. Nature. 2023;616:123–131.

39. Lin AE, Bapat AC, Xiao L, et al. Clonal Hematopoiesis of Indeterminate Potential With Loss of *Tet2* Enhances Risk for Atrial Fibrillation Through *Nlrp3* Inflammasome Activation. Circulation. 2024;149:1419–1434.

40. Saadatagah S, Naderian M, Uddin M, et al. Atrial Fibrillation and Clonal Hematopoiesis in TET2 and ASXL1. JAMA Cardiol. 2024;9:497–506.

41. Ahn H-J, An HY, Ryu G, et al. Clonal haematopoiesis of indeterminate potential and atrial fibrillation: an east Asian cohort study. Eur Heart J. 2024;45:778–790.

42. Kallai A, Ungvari A, Csaban D, et al. Clonal hematopoiesis of indeterminate potential (CHIP) in cerebromicrovascular aging: implications for vascular contributions to cognitive impairment and dementia (VCID). GeroScience. 2025;47:2739–2775.

43. Marnell CS, Bick A, Natarajan P. Clonal hematopoiesis of indeterminate potential (CHIP): Linking somatic mutations, hematopoiesis, chronic inflammation and cardiovascular disease. Journal of Molecular and Cellular Cardiology. 2021;161:98–105.

44. Straub RH. The complex role of estrogens in inflammation. Endocr Rev. 2007;28:521–574.

45. Choi HJ, Lee AJ, Kang KS, Song JH, Zhu BT. 4-Hydroxyestrone, an Endogenous Estrogen Metabolite, Can Strongly Protect Neuronal Cells Against Oxidative Damage. Sci Rep. 2020;10:7283.

46. Xing D, Nozell S, Chen Y-F, Hage F, Oparil S. Estrogen and Mechanisms of Vascular Protection. ATVB. 2009;29:289–295.

47. Zarezadeh Mehrabadi A, Shahba F, Khorramdelazad H, et al. Interleukin-1 receptor accessory protein (IL-1RAP): A magic bullet candidate for immunotherapy of human malignancies. Critical Reviews in Oncology/Hematology. 2024;193:104200.

48. Svensson EC, Madar A, Campbell CD, et al. TET2-Driven Clonal Hematopoiesis and Response to Canakinumab: An Exploratory Analysis of the CANTOS Randomized Clinical Trial. JAMA Cardiol. 2022;7:521–528.

49. Woo J, Lu D, Lewandowski A, et al. Effects of IL-1β inhibition on anemia and clonal hematopoiesis in the randomized CANTOS trial. Blood Advances. 2023;7:7471–7484.

50. Woo J, Zhai T, Yang F, et al. Effect of Clonal Hematopoiesis Mutations and Canakinumab Treatment on Incidence of Solid Tumors in the CANTOS Randomized Clinical Trial. Cancer Prev Res (Phila). 2024;17:429–436.

51. Corty R, Pershad Y, Vlasschaert C, et al. Detection of clonal hematopoiesis of indeterminate potential via genome or exome sequencing profoundly underestimates disease associations. medRxiv. 2025:2025.08.11.25333294.

